# HIV, HPV, and Oral Health in Tanzania: A scoping review

**DOI:** 10.1101/2025.02.05.25321725

**Authors:** Kalipa Gedion, Elizabeth Blackwood, Judith Mwobobia, Innocent Semali, Mainen Julius Moshi, Sira Owibingire, Richard O Mwaiswelo, Yohana Mashalla, Guido Ferrari, John Bartlett, Nosayaba Osazuwa-Peters

## Abstract

**Background:** There is an increased risk of human papillomavirus (HPV)-associated infections and malignancies among people living with HIV (PLHIV). However, there is limited literature exploring the intersection of HPV, HIV, and oral health in Tanzania and across sub-Saharan Africa. We reviewed the existing literature on the intersection of HIV, HPV, and oral health in Tanzania.

**Methods:** This was a scoping review with the search of key words representing HIV, HPV, oral health, and Tanzania. Since there were no studies that explored the intersection of HIV, HPV, and oral health in Tanzania, the search extended to include studies with the intersection between oral health and either HIV or HPV in Tanzania.

**Findings:** 44 studies were eligible for analysis. Only one of them explored the relationship between HPV and oral health, where 4 (6%) of adolescent schoolgirls were detected with HPV-DNA and the paper hinted at the possibility of HPV autoinoculation. There were no articles linking HPV vaccination and oral health. The remaining 43 (98%) studies explored the relationship between HIV and oral health. There has been an increase in oral manifestations in PLHIV in the last two decades, and highly active antiretroviral therapy has been protective against oropharyngeal candidiasis but had no significance on head and neck cancer. Single-dose fluconazole and 35% herbal antifungals were identified to be effective in treating oral candidiasis. No recent studies explored the different facets of dental care among PLHIV.

**Interpretation:** There are no studies exploring the intersection of HIV, HPV, and oral health in Tanzania. Future studies are needed to determine the burden and barriers of HPV-associated oral manifestations among PLHIV in Tanzania and across Sub-Saharan Africa.

## Introduction

At least two-thirds of the 39 million people living with HIV (PLHIV) globally reside in sub-Saharan Africa, with the highest reported AIDS-related mortality of 380,000 people annually.(1) HIV is also associated with increased risk of infection by the human papillomavirus (HPV), and evidence suggests that in sub-Saharan Africa, up to 35% of women living with HIV (WLHIV) and 39% of men living with HIV (MLHIV) have acquired high-risk HPV (hr-HPV) infection.(2,3) In Tanzania, 12% of WLHIV and 25.7% of MLHIV report hr-HPV infection; this rate of infection by hr-HPV seems to be significantly increased in PLHIV, compared to non-PLHIV.(4,5) The prevalence of HIV in Tanzania has reduced from 5.3% in 2011 to 4.5% in 2021, partly due to vigorous HIV/AIDS prevention programs. The country has achieved the 95-95-95 targets: at least 95% were diagnosed with HIV, 95% were on ART medications, and 95% attained viral load suppression.(6) This has led to an improved life expectancy for PLHIV. However, this longer-term survival of PLHIV has also made cancer prevention an imperative as cancer has emerged a competing cause of mortality for PLHIV.(7) Of these cancers, cervical cancer is the leading cancer among women and HPV-related cancers in Tanzania.(8,9)

Globally, the link between HPV and cervical and other anogenital cancers have been much more explored compared to HPV’s association with head and neck cancer..(10–12) In addition to head and neck cancer, HPV is also associated with oropharyngeal candidiasis in the head and neck region.(11,12) These conditions are significantly more common in individuals with a history of HIV.(13) Additionally, given the anatomic location, oropharyngeal candidiasis and HPV-associated oropharyngeal cancer both have oral health implications, furthering broadening the link between oral health and systemic health.(14–18) However, the association of PLHIV with HPV-related oral manifestations remains largely unexplored in sub-Saharan Africa.

Due to the high burden of morbidity and mortality associated to cervical cancer (10,241 diagnosed women and 6525 deaths annually), Tanzania introduced a national HPV vaccination campaign for 14-year-old girls nationally in 2018.(19) The current vaccination campaign covers young girls only, resulting in potential gaps in coverage for other vulnerable individuals, including those PLHIV and others outside of this country-specific age of eligibility. The current campaign’s coverage is in contrast with the Centers for Disease and Control (CDC), recommending the vaccination of males and females aged 11 to 26 years, and a recent expansion of the age of eligibility in the United States to 27-45 years, based on a patient-physician shared decision making model.(20,21) This underscores a need to improve the coverage of the HPV vaccination protocol in Tanzania and across sub-Saharan Africa.

Given the limitation of resource availability in Tanzania and other low-middle-income countries (LMICs) in sub-Saharan Africa, it is critical to explore the complex interplay between HIV, HPV, and oral health. Individuals with HIV have 2-3 higher odds of contracting oral HPV infections thane HIV-uninfected population.(22) HIV individuals were more likely to have an hr-HPV genotype and persistent uncleared infection.(23) Furthermore, HIV-infected individuals had an increased risk of acquiring HPV-associated and unassociated head and neck cancer.(7,23) The association between HIV and HPV-associated head and neck cancer has not been thoroughly explored in sub-Saharan Africa. A recent study in India reported that lower HPV oral infections were linked to HPV vaccination.(24) While sub-Saharan Africa has studies on HPV vaccination, none have explored its connection to oral manifestations.(25,26) Understanding the interplay of HIV, HPV, and oral health in Tanzania is important for measuring the burden, defining the barriers, and formulating interventions. This study aimed to fill this gap by conducting a scoping review of the intersection of HIV, HPV, and oral health in Tanzania.

## Methods

### Search Strategy

The search was developed and conducted by a professional medical librarian (E.B.) in consultation with the author team (K.G. and N.O.P.), and included a mix of keywords and subject headings representing oral health concerns, HPV, HIV, and Tanzania. The searches were independently peer reviewed by another librarian using a modified PRESS Checklist (McGowan).(27)

Searches were conducted in MEDLINE via PubMed, Embase via Elsevier, Web of Science via Clarivate, and Global Index Medicus via the World Health Organization, and CABI Global Health via EBSCOhost. The searches were executed on February 9, 2024 and found 423 unique citations. Complete reproducible search strategies, including search filters, for all databases are detailed in the Supplementary Materials. All citations were imported into Covidence, a systematic review screening software, which also de-duplicated the citations.

### Criteria for Inclusion

To be included in the study, papers had to document an intersection between oral health and HIV or HPV. Additionally, it was required that study populations were based in Tanzania only.

### Criteria for Exclusion

Authors excluded papers that fell in the wrong geographic region (outside of Tanzania) or were of mixed settings (included Tanzania, but also included other regions). The authors also excluded papers that did not contain empirical data, did not explore the intersection of oral health and either HIV or HPV, or were of the wrong study type. Specifically, systematic reviews, scoping reviews, abstracts, case reports, and editorials. Additionally, papers that were not published in English were excluded from the study; however, the authors note that this is a limitation of the current project and offers room for further study. Two coauthors (K.G. and J.M.) reviewed extracted literature, and all disagreements were resolved by N.O.P.

## Results

### Study Selection

A total of 840 records were identified from database searches, and 423 titles and abstracts were screened after duplicates were identified and removed. Following selection criteria, 44 studies from Tanzania were included in our final analysis (see Fig 1). Most of the studies were cross-sectional or cohort studies; however, there were also a few randomized controlled trials, and one qualitative study included (see appendix). There is a profound gap in the knowledge of HIV, HPV, and oral health, as we found no studies in the intersection of HIV, HPV, and oral health in Tanzania (see Fig 2).

**Fig 1:**
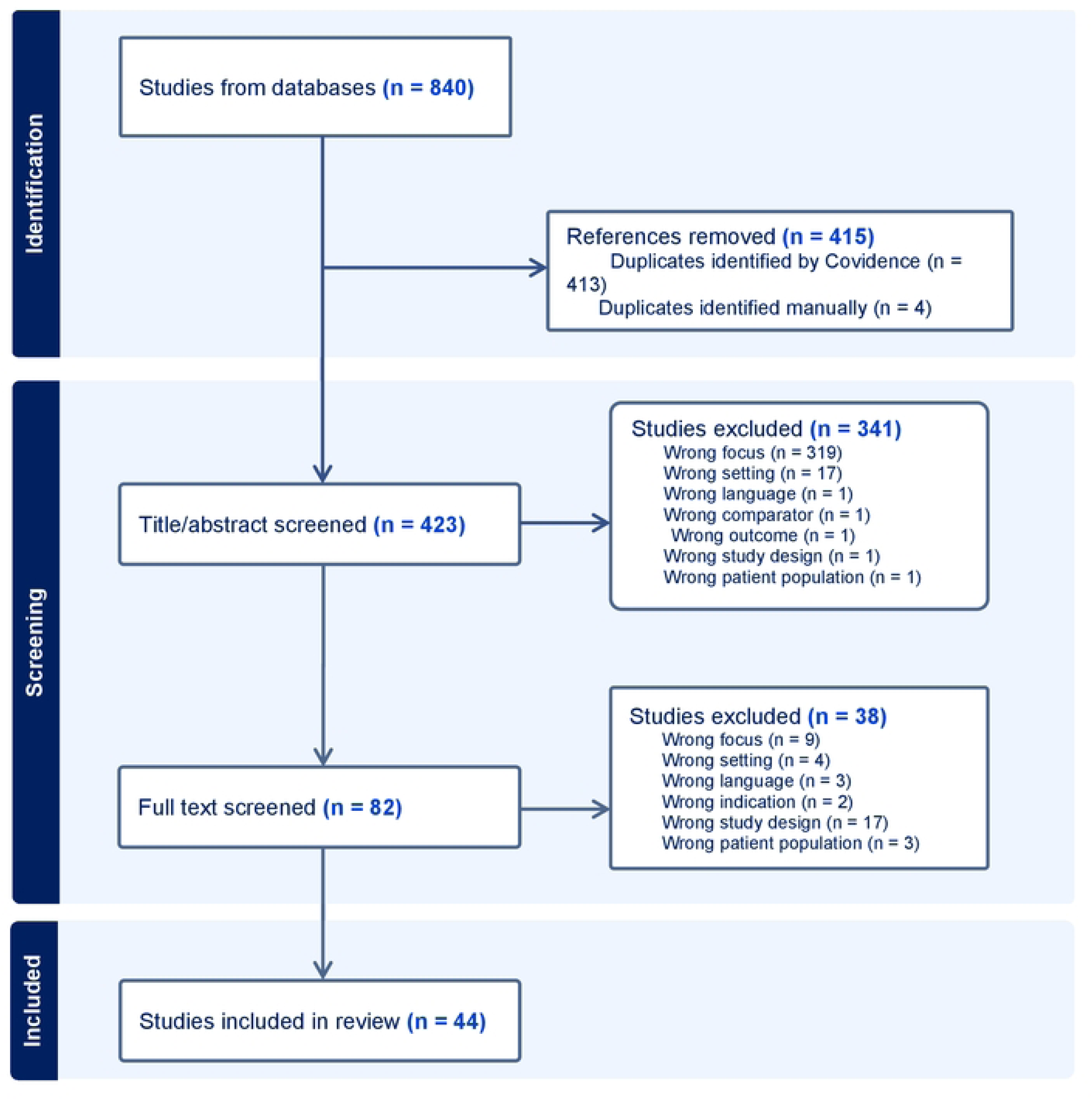
Study flow chart showing inclusion and exclusion criteria.

**Fig 2.**
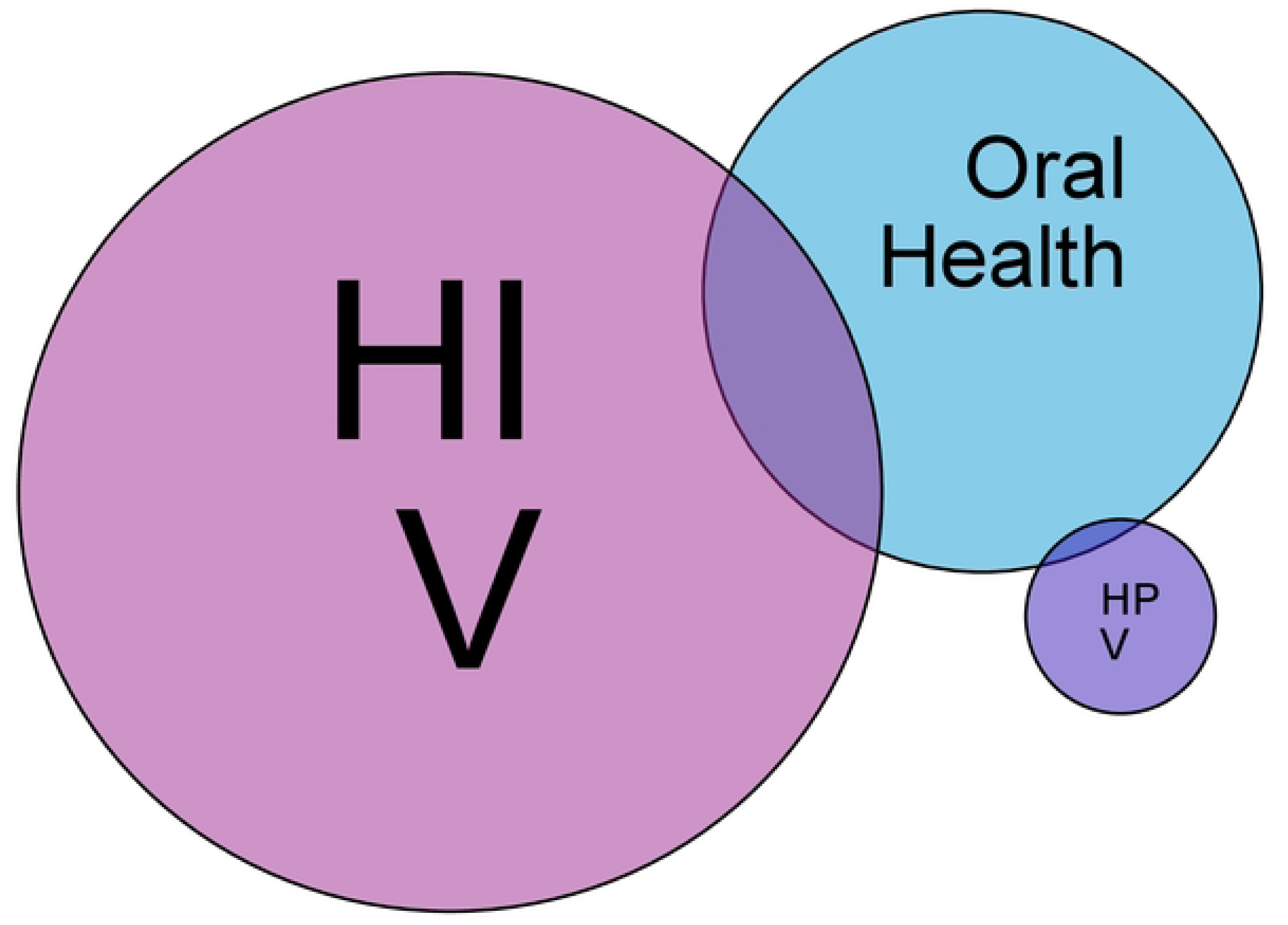
Venn diagram highlighting the intersection of the 44 studies. NB: Color should be used for any figures in print.

### HPV and oral health

To generalize our search to attain any relationship of the HPV and oral health in Tanzania, only one paper researched the presence of HPV in oral, vaginal, and toilet surfaces in adolescent girls who reported a sexual debut.(28) Six percent of 63 girls had HPV DNA detected in the oral region, higher than 4.5% of normal adult population reported in a systematic review.(28) 82% of these adolescent girls reported to not perform fellatio, and 1 out of 41 who agreed to test for HIV was positive.(28) Findings also suggested that, because of the similarities of genotypes between the sites, there was a possibility of autoinoculation.(28) Studies regarding HPV autoinoculation have mixed findings and requires further analysis on its impact on the oral health of PLHIV.(29,30)

Furthermore, there are no studies exploring the effect of HPV vaccination on oral health in Tanzania. A 2019 study in the U.S. revealed a similar and plateaued HPV antibody levels following three doses of HPV vaccine for both MLHIV and non-MLHIV.(31) The study strongly supports the HPV vaccination for the prevention of infection and cancer in the oral cavities of men.(31) More studies are needed to evaluate the effectiveness of HPV vaccines in oral health of PLHIV for all sexes. While PLHIV on HAART (highly active antiretroviral therapy) without vaccination are developing less HPV oral lesions, they are still prone to HIV oropharyngeal cancers, and there is contradictory evidence of the therapy’s protection of cervical cancer. (7,32–36) Research is necessary to identify why HPV oral cancers are persistent in PLHIV on HAART.

### HIV and oral health

Studies exploring the link of HIV and oral health revealed that the increase of PLHIV with at least one oral manifestation was from 10% to 34% from 2000 to 2020.(37–42) This tripled trend of PLHIV is alarming and calls for further study and immediate intervention. Other studies elsewhere revealed oral manifestations up to 50% in PLHIV and 80% in PLWA in 2019.(43) These lesions were associated with HIV clinical stage and smoking.(39) Majority (59 - 90%) of PLHIV had sound awareness of oral manifestations, increasing with education status.(37,44) However, less than half has sought medical attention.(37) This underscores a need to measure the current awareness and health-seeking behavior of PLHIV to form well-informed interventions. Moreover, PLHIV on HAART had a significantly lower risk of oral lesions and candidiasis.(39,45) This coincides with other studies elsewhere.(46,47)

Oropharyngeal candidiasis (OPC) presented to be a major predictor of HIV in oral manifestations, with a prevalence ranging from 4 to 42%, where up to 61% in children and 50% in newly diagnosed with HIV.(37–42,48–52) Other studies elsewhere have a wide prevalence from 17% to 75%.(53–55) OPC was linked to HIV status, low CD4 counts, HIV mortality, and weight loss, ART-naivety, which correlates with other countries.(39–41,48,56–61) Multivitamins had shown to be a protective factor of OPC and lower concentration (<10ng/ml) of Vitamin D was a risk factor.(62–64) The most common agent for OPC was candida albicans, but 6.5% were the remaining non-albicans candida – more common in PLHIV.(65) Lastly, there was no significance of oral thrush and completion of T.B. treatment in HIV mortality because OPC is an independent predictor of HIV mortality.(66,67)

There was no difference in oral herpes detection between compromised vs uncompromised HIV children, with a prevalence of 3.2% in PLHIV.(40,68) Some of other oral manifestations were angular cheilitis (5% - 7%), oral hairy leukoplakia (1% - 36%), followed by Kaposi sarcoma (>1% - 4%) in PLHIV.(37,39–41) Recent studies elsewhere reported a higher prevalence of these manifestations, including oral herpes (4 - 20%), angular cheilitis (3% - 20%), and oral hairy leukoplakia (8 - 18%).(55,69,70) There is a need for a follow-up study of the current situation of oral manifestations in Tanzania.

Eighty-one percent of HIV children tested positive for pneumonia nasopharyngeal swabs, but it is less frequent than HIV-uninfected children.(71,72) Conversely, S. aureus was more common in CHIV but diminished over time.(73)

### HIV-related oral malignancies

In 2000, two out of 125 PLWA (people living with AIDS) had oral carcinoma.(38) Of all non-AIDS-defining cancers, 81.5% of them had HNC in Tanzania, and they make up 7.5% of all HNC cases in the general population.(74) Almost half of the HNC of PLHIV originated from the oropharynx, followed by the nasal cavity 27.2%, and hypopharynx/ larynx/ trachea 9.9%.(74) A US study revealed that PLHIV with HNSCC had a lower survival rate (HR:1.98) and worse outcomes than those without HIV.(75) The tumors of PLHIV with HNSCC had lower intratumoral CD8 infiltration despite these patients being immunocompetent (normal CD4 counts and viral load).(75) Furthermore, over half of PLHIV with HNC were diagnosed at the advanced malignancy stage in Tanzania, similar to the U.S.(75,76) We have limited data on the prognosis of PLHIV with HNC and require early cancer detection protocols.

In Tanzania, 11% of Oral Kaposi Sarcoma (OKS) cases had 69.7% PLHIV.(76) Another study showed that 0.5% −3.5% of PLHIV had OKS manifestation.(39) This is quite lower than Brazil’s more recently reported prevalence of 36%.(77) In Tanzania, OKS was associated with HIV, advanced histological staging, decreased CD4 count, increased tumor burden in men, and increased frequency in females.(76) Another study in Brazil presented similar CD4 findings but also showed that 22.8% of PLWA with K.S. had oral lesions, but frequently in young black men.(77) Recent studies are required to determine the prevalence and risk factors of OKS in Tanzania.

Additionally, while HAART proved to be protective against oral lesions and cervical cancer in Tanzania, there was no statistical significance of the therapy on HNC.(78) This finding aligns with some studies in other countries.(46,79,80)

### Traditional and pharmaceutical antifungal treatments

This review found studies regarding candida treatment in oral health among PLHIV in Tanzania. Of medicinal plants, 24 - 25% were identified by herbal practitioners for treating candidiasis, and 35% of the known medicinal plants were effective.(81,82) Uganda’s herbalists treat oral candidiasis using different herbs from those in Tanzania, and Iranians use essential oils to treat fluconazole-resistant candida infections of PLHIV.(83) This indicates the diversity of effective herbal options across the globe. There underlines a great need to document these indigenous practices for further exploration.

Candida in Tanzania has a high susceptibility to antifungal agents with minimal resistance.(84) Another study in 2008 revealed the similar effectiveness of a single dose vs 2-week fluconazole administration for oropharyngeal candidiasis, similar to other studies.(85–87) A single dose was considered cost-effective in resource-constrained settings and with higher adherence. Still, this treatment has not been adopted in the Standard Treatment Guidelines and other international treatment protocols.(85,88,89) More studies are warranted to determine the effectiveness and adherence of a single dosage to its adoption to treatment guidelines.

### Dental practice

Three studies in Tanzania identified no difference in dental caries, safety, or post-extraction complications between PLHIV and non-PHIV in dental care.(61,90,91) Other studies elsewhere pointed out that dental caries was more prevalent in PLHIV, with 42% to 83%, unlike in Tanzania.(70) An old study in 1992 shared findings that dental workers were unfamiliar with oral manifestations of PLHIV.(92) No follow-up study has been conducted since. Furthermore, studies elsewhere explored continual stigma to PLHIV, rapid testing willingness, medico-legal concerns, risk assessment screening, and varied barriers in dental settings.(93–97) More studies are needed to explore different facets of dental care among PLHIV.

## Discussion

There exists an extensive knowledge gap in the intersection of HPV, HIV, and oral health in Tanzania. No studies are mapping this intersection to determine the burden of the problem. In developed countries, there is a doubled prevalence of HPV oral manifestations in PLHIV versus non-PLHIV. However, there is no data to compare in sub-Saharan Africa.(7,98) Tanzania’s health systems have been strengthening to combat infectious diseases, but because of the epidemiologic transition, there is an increased burden of NCDs.(99,100) In the U.S., HPV-associated oropharyngeal cancer has surpassed cervical cancer as the most common HPV-related cancer.(11) Without immediate action to understand and manage the burden of HPV oral malignancies in Tanzania, health systems will be left unprepared and overwhelmed to tackle HPV oral cancers. There is no determined prevalence of PLHIV with HPV oral manifestations or a recent prevalence of PLHIV with any oral manifestations in Tanzania. No studies identified the risk factors that may contribute to HPV oral manifestations in PLHIV. Studies have shown that oral sex is a primary predictor of hr-HPV oral detection and cancer, especially in men.(101–104) Oral sex practice is common in developed countries, but it is increasingly growing (47%) among university students and adults in sub-Saharan Africa, especially in men.(105) In Nigeria, the odds of oral sex were increased among PLHIV, men who have sex with other men (MSM), and multiple sexual partners but also associated with a higher oropharyngeal STI prevalence.(106) While there is limited data on oral sex practice in Tanzania, neighboring countries hint at an upward trend. This risk factor can likely lead to the increase of HPV oral transmission in both sexes. Furthermore, men are prone to acquiring the oral HPV virus and are unprotected by the current HPV guidelines in Tanzania. They are at an even greater risk of exposure and malignancy.(19,107) It is paramount to gather evidence to explain the intersection to form effective interventions. We call to research the prevalence, incidence, and risk factors of HPV oral manifestations among PLHIV in Tanzania.

In 2018, the government of Tanzania implemented the HPV vaccination of up to 14-year-old girls.(19) The coverage is limited compared to the recommendations of the CDC and WHO. CDC recommends vaccinating both men and women from the age of 11 to 26.(21) WHO recommends the vaccination of girls till 14 to the catch-up age of 19, but also immunocompromised or HIV patients of any gender or age.(28) WHO also recommends vaccinating older women, men, or MSM if it is feasible and affordable.(28) Developed countries have been experiencing an increase in HPV oropharyngeal cancers in all sexes but higher in men.(107,108) It is likely that Tanzania will follow suit because of the epidemiological shift. It is paramount to extend the vaccination coverage to MLHIV as studies have shown to protect them from HPV infection and cancer.(31) Furthermore, an alarming 2023 study reported that WLHIV in Tanzania had a 6-fold higher prevalence of HPV 52 than non-WLHIV.(4) This high-moderate HPV genotype is not covered by the quadrivalent vaccine offered in Tanzania because of its cost-effectiveness.(10,109) There is a lack of evidence of HPV 52 persistence and progression to cancer. Urgent attention is needed for further research on selecting an appropriate broad HPV vaccine for the Tanzanian population and reevaluating the vaccination coverage.

It is worth noting that our study had limitations. Our study included papers with study designs with empirical evidence, excluding reviews and case reports. It limited the exploration of the subject matter. The review also excluded studies that were not geographically bound to Tanzania solely. The intention was to minimize the lack of generalization of our data. Furthermore, the review included recent and older evidence, regardless of the publication date, for their valuable insights. Because of the long time range, some findings might not reflect the country’s current situation. It also excluded a few studies without full articles despite the efforts for retrieval. This may have led to selection bias and affected the findings. Lastly, the quality of the included studies differed in methodological approaches and flaws. This may have introduced bias and affected the conclusions.

### Global health implications of study

This scoping review identified an important gap in the intersection of HIV, HPV, and oral health, as we found no studies throughout the Tanzanian literature. This limitation, or paucity of data that interlinks oral health, HIV and HPV is also evident throughout the sub-Saharan Africa region, where HIV is endemic, and with an increasing burden of HPV-associated malignancies. In contrast, data indicate that 23% of PLHIV in the US has oral HPV infection, more than double the prevalence of non-PLHIV. MLHIV had a higher prevalence than WLHIV, and oral HPV among PLHIV was linked to oral sex partners and immunodeficiency.(110) Regarding detection through oral washes, immunocompetent PLHIV were likely to have low-risk HPV types, while immunocompromised PLHIV had high-risk HPV types, increasing their risk of HPV-associated oropharyngeal cancer.(111) The U.S. has an increasing trend of HPV-associated oropharyngeal cancer in PLHIV from 6.8 - 11.44 cases per 100,000 years from 1996 to 2009.(112) Due to the epidemiologic shift of increasing NCD burden, there could be an increase in the burden of HPV-associated malignancies in Tanzania in the future.(113,114) However, the incidence of such cases is unknown. Another U.S. study revealed that PLHIV with HPV-oropharyngeal tumors had a worse prognosis and lower survival. However, it was a small-scale study and presented contrasting evidence from previous studies.(75) While HPV HNSCC (HPV head and neck squamous cell carcinoma) tends to have more favorable prognoses than non-HPV HNSCC, the phenomenon is unclear among PLHIV and warrants further studies.(75,115)

Given the burden of disease, the fact that there is little or no data linking HIV, HPV and oral health in Tanzania suggests that there is a critical need for more research infrastructure and capacity building in this area of research.

## Conclusions

There are no studies exploring the intersection of HIV, HPV, and oral health in Tanzania. It is important to map the intersection and determine the burden and barriers of HPV-oral manifestations among PLHIV in Tanzania. This will inform stakeholders in developing and improving evidence-based protocols. Further research in this intersection will fill the knowledge gap in sub-Saharan Africa and globally.

## Data Availability

N/A

## Acknowledgements

Dr. Barlett is supported by NIH awards TW D43 009595 and AI P30 064518. Dr. Osazuwa-Peters is supported by NIH award R01DE032216

## Appendix: Table

**Table 1:** Characteristics and findings of the 44 articles.

| Surname, year | Aim of study | Study design | Population description | Related findings of the intersection |
| --- | --- | --- | --- | --- |
| Houlihan 2019(28) | To describe the HPV DNA prevalence in genital and non-genital sites and in the bathroom of unvaccinated adolescent girls, and examine genotype concordance between sites | Cross sectional study | 65 Unvaccinated girls aged 16-18 years in Mwanza, who reported ever having had sex | Six percent of the 63 adequate oral samples had detectable HPV. |
| Zuckerman 2015 | To evaluate longitudinal oral HSV reactivation in HIV-positive and negative children. | Cohort study | 20 HIV positive ART-naive and 10 HIV negative aged 3-12 years | There was no difference in HSV detection between uncompromised HIV-infected and HIV-uninfected children.<br>HIV-infected children who reported having oral sores have a high rate of HSV detection. |
| Villamor 2005(56) | To examine the effect of vitamin supplements on wasting in HIV-infected women and to assess the effects of sociodemographic characteristics, morbidity events, and immunologic progression on the risk of wasting. | Randomised controlled trial | 1078 HIV infected women in Tanzania | Oral ulcers and thrush were significantly related to the increased risk of wasting. |
| Tillekeratne 2009(57) | To identify the components of an effective Community home-based care (CHBC) program | Cohort study | 257 HIV infected adults | Oral thrush and dysphagia were associated with HIV mortality in 2 years. |
| Sudfeld 2013(58) | To assess associations of body mass index (BMI; in kg/m<sup>2</sup>) at antiretroviral therapy (ART) initiation and weight change after 1 mo of treatment with mortality, morbidity, and CD4 T cell reconstitution | Cohort study | 3389 Tanzanian adults initiating ART enrolled in a multivitamin trial | Weight loss of HIV patients at 1 month of ART was associated with oral thrush. |
| Sudfeld 2015(62) | To prospectively assess the association of vitamin D status with mortality, morbidity, and growth during the first 2 y of life. | Cohort study | 53 HIV-infected and 948 HIV-exposed Tanzanian infants | Among HIV-exposed infants, lower concentrations (<10 ng/mL) of Vitamin D were associated with oral candidiasis and wasting. There was no statistically significant association between lower concentrations of Vitamin D and HIV mortality. |
| Ngasala 2016(48) | To determine the prevalence of oral candida infection in HIV positive patients and investigate the relationship between oral manifestations and the level of immunosuppression. | Cross sectional study | 314 HIV patients on ARV and attending hospital for care and treatment | The prevalence of oral candida was 42.0% in HIV individuals, more than half being in the age group 6-17 years. A significantly higher prevalence of candida infection (66.7%) was observed among patients with <200 cells/ $\mu$ l than in those with 200-500 cells/ $\mu$ l or >500 cells/ $\mu$ l. The prevalence of oral candida infection was significantly higher in patients with CD4+ cell counts of less than 200 cells/ $\mu$ l. |
| Mwangosi 2011(37) | To assess the magnitude of and extent of awareness on oral manifestations among PLWHA attending counseling and treatment centers (CTC) in Iringa Municipality in south-western Tanzania. | Cross sectional study | 200 PLWHA | 23.5% of the PLWHAs had at least one oral manifestation, with statistically significant differences across age groups.<br>Clinical manifestations observed were mucosal ulcerations with or without severe periodontal lesions (7.0%), angular cheilitis (7.0%), oral thrush (6.5%), Kaposi's sarcoma (1.5%), and hairy leukoplakia (1.0%).<br>The majority (89.5%) of PLWHA had sound awareness of clinical oral manifestations with a significant statistical difference by educational status. |
| Mwangosi 2012(49) | To assess the prevalences and patterns of oral lesions occurring in human immunodeficiency virus (HIV) and acquired immune deficiency syndrome (AIDS) | Cross sectional study | 200 HIV subjects | 58.5% were aware of predispositions towards the occurrence of oral lesions such as oral candidiasis (60.0%) in HIV/ AIDS an 72.0% were aware that the lesions are treatable.<br>Some participants reported occurrences of oral thrush (22.5%) and lip ulcerations (28.5%), although only 47.0% of these had sought medical advice.<br>29.0% of participants had at least one oral lesion associated with HIV/AIDS: 11.5% for herpes simplex, 7.5% for oral candidiasis, 4.0% for oral hairy leukoplakia, 3.5% for Kaposi's sarcoma; 1.5% for dry mouth; 0.5% for<br>angular cheilitis, and 0.5% for acute necrotizing ulcerative gingivitis. Herpes simplex and Kaposi's sarcoma were more frequently observed in males (56.5% and 71.4%, respectively), whereas oral candidiasis and dry mouth were observed more often in females (86.7% and 66.7%, respectively). |
| Mushi 2016(65) | To compare the oral colonization of non-albicans candida spp. between HIV and non-HIV infected individuals in Mwanza, Tanzania. | Cross sectional study | 351 HIV-infected and 639 non-HIV infected in Mwanza without clinical signs of oral candidiasis | Candida albicans was the commonest spp. detected in both groups 119 (n = 351, 33.90%) of HIV-infected and 146 (n = 639, 22.9%) of non-HIV infected population (p = 0.023).<br>NAC spp. carriage was detected in 64 (n = 990, 6.46%) of the study participants. Of HIV-infected patients, 36/351 (10.3%) carried NAC spp., as compared to only 28/639 (4.4%) among non-HIV infected (P = 0.0003).<br>Only one HIV infected participant was colonized by two NAC spp.<br>The median CD4 cell count among HIV-infected patients colonized by non-albicans Candida spp. was 251 (IQR 170-380) cells/ml, while for those not colonized by NAC spp. was 473 (IQR 304-673.5) cells/ml, P < 0.001.<br>Of 191 HIV-infected individuals without history of antibiotic use, 15 (7.85%) were colonized by NAC spp., as compared to 26 (4.3%) of 606 HIV-uninfected individuals without a history of antibiotic use p = 0.026 |
| Mugusi 2012(66) | To describe risk factors for mortality and clinical characteristics of HIV-infected patients with and without tuberculosis (T.B.) coinfection | Cohort study | 255 HIV infected patients without active T.B. and 231 HIV infected patients with active T.B., all with CD4 less than 200 | There was no difference between oral candidiasis infectivity among those with or without T.B. Oral candidiasis is a predictor of mortality of HIV patients (regardless of the status of T.B.). |
| Mugusi 2009(67) | To investigate factors associated with mortality, including patients' HIV serostatus, CD4 cell count, laboratory, nutritional and demographic characteristics in AFB smear positive pulmonary T.B. patients. | Randomized controlled trial | 887 TB positive patients aged 18-65 receiving 8 months anti-TB treatment | Oral thrush was associated with mortality of HIV-infected patients. The association still exists after 8 months of treatment of T.B. Causes of death could be due to other conditions apart from T.B., likely HIV-related causes. |
| Miller 1995(59) | To directly compare previously proposed modifications of the original WHO CCD among medical inpatients in a large urban teaching hospital in Tanzania | Cross sectional study | 223 inpatients in Muhimbili aged 16 and above | Oral candidiasis and lymphadenopathy as best HIV predictor. |
| Mehta 2011(63) | To examine the relationship between low vitamin D status and HIV-related complications, | Randomized controlled trial | 884 HIV infected women in Tanzania | Low Vitamin D is associated with the complication of oral thrush and reported mouth/throat ulcers. |
| Matee 2000(38) | To determine the association, if any, between the presence of oral lesions and clinical | Cross sectional study | 192 HIV-infected individuals not receiving treatment and 156 individuals confirmed to | Intra-oral lesions were seen among 7.7% of the HIV-seronegative, 10.4% of the HIV-seropositive, and 36.8% of the AIDS groups, respectively. |
|  | and immunological status of untreated HIV-infected adults in Tanzania. |  | be HIV-seronegative acted as a control group | Enlarged parotid glands were seen in 20% of the AIDS patients, 11.9% of the HIV-seropositive, and 5.1% of the HIV seronegative.<br>Enlargement of submandibular salivary glands was seen in 29.6% of the AIDS patients, 31.3% of the HIV-seropositive compared with 14.7% among the HIV-seronegative.<br>The association of oral lesions with the clinical stage of HIV infection and to a lesser extent peripheral CD41 T cell count does suggest that these lesions could be used as additional markers of immunosuppression and AIDS. (OR higher among seropositive, then decreasing CD4 counts, and total lymphocyte counts)<br>Intra-oral lesions occurred in 36.8% of the AIDS patients, which is approximately 3.5 times the frequency among the HIV-seropositive (10.4%) and 4.8 times the frequency among the HIV-seronegative individuals (7.7%).<br>Oral candidiasis and hairy leukoplakia were the most frequent lesion.<br>Carcinoma only seen in AIDS patients 2(1.6%) of 125. |
| Masoza 2022<sup>27</sup> | To determine the prevalence, clinical characteristics and outcome of HIV-infected children admitted at Bugando Medical Centre (BMC) after active provision of PITC services. | Cross sectional study | 525 inpatient children | Factors independently associated with oral thrush (OR 20.06).<br>61.2% of HIV infected had oral thrush while 3.6% of uninfected had oral thrush. |
| Kinabo 2013(73) | To estimate the prevalence of nasopharyngeal bacterial colonization (NPBC) patterns in young Tanzanian HIV-exposed infants and to analyze the influence of maternal NPBC and of the infant's HIV status on the NPBC pattern. | Cohort study | 338 HIV exposed infants | In HIV-uninfected infants, colonization with *S. pneumoniae* was more frequent at 6 weeks, 3 and 6 months of age, than in HIV-infected infants while colonization with *S. aureus* diminished over time and was more common in HIV-infected infants. Co-colonization of *S. pneumoniae* with *H. influenzae* or *M. catarrhalis* was mostly noticed in HIV- infected infants. Maternal *S. aureus* colonization was a risk factor for colonization in HIV-infected infants. |
| Kapiga 1994(51) | To identify risk factors for HIV infection among women not known to be members of high-risk groups in Dar es Salaam, Tanzania and assess associations between contraceptive use and HIV infection | Cross sectional study | 2285 women, not in key and vulnerable groups | Candidiasis found in 15% of HIV-infected women. |
| Kahabuka 2007(44) | To investigate the awareness of the oral manifestations of HIV/AIDS and general issues about HIV and AIDS among people living with HIV | Cross sectional study | 187 PLHIV in Dar es Salaam | 13.4% of the participants were completely unaware of the oral manifestations of HIV/AIDS. Participants were relatively well aware of the different types of oral manifestations (e.g., oral ulcers 87%, oral candidiasis 84%) while their knowledge of the management of |
|  | (PLHIV) in Dar es Salaam, Tanzania |  |  | specific oral manifestations and the problems associated with oral manifestations was more limited. |
| Hamza 2006(39) | To compare the prevalence and types of HIV-related oral lesions between children and adult Tanzanian patients on HAART with those not on HAART and to relate the occurrence of the lesions with anti-HIV drug regimen, clinical stage of HIV disease and CD4+ cell count | Cross sectional study | 532 HIV infected patients, 51 children and 481 adults, 165 males and 367 females. Children were aged 2 to 17 years and adults 18 and 67 year | HIV-associated oral lesions were observed in 39.5% of patients. Oral candidiasis was the most common (23.5%), followed by mucosal hyperpigmentation (4.7%), parotid enlargement (3.9%), and oral Kaposi's sarcoma (3.2%). In children, parotid enlargement (19.6%) was more common than oral candidiasis 11.8%. There was a significant difference in the occurrence of oral candidiasis and parotid enlargement between children and adults.<br>Adult patients on HAART had a significantly lower risk of oral lesions, oral candidiasis, and oral hairy leukoplakia. There was no significant reduction in the occurrence of oral lesions in children on HAART.<br>There was also a significant association between the presence of oral lesions and CD4+ cell count < 200 cell/mm<sup>3</sup> and with WHO clinical stage .<br>Oral lesions were also associated with tobacco smoking. |
| Hamza 2008(84) | To determine the species distribution of Candida isolates obtained from Tanzanian HIV infected | Cross sectional study | 292 HIV infected patients with oropharyngeal candidiasis at MNH | No statistical difference between primary and recurrent oropharyngeal candidiasis.<br>Recurrent oropharyngeal candidiasis |
|  | patients with primary and recurrent OPC. |  |  | and previous antifungal therapy significantly correlated with reduced susceptibility to azoles antifungal agents.<br>C. albicans most frequently isolated species from oropharyngeal candidiasis.<br>Oral yeast from Tanzania had a high level of susceptibility to antifungal agents. |
| Fawzi 2004(64) | To examine the effects of daily supplements of vitamin A (preformed vitamin A and beta carotene), multivitamins (vitamins B, C, and E), or both on progression of HIV disease, using survival models | Randomized controlled trial | 1078 pregnant women infected with HIV | Supplementation with multivitamins reduced the incidence of oral thrush, oral ulcers, difficulty in swallowing, and esophageal candidiasis.<br>Micronutrients may protect the integrity of oral and gastrointestinal epithelia and enhance local and systemic immunity. |
| Faini 2015(60) | To determine the incidence and prevalence of fungal infections in Tanzania | Other |  | Oral candidiasis was the most common clinical presentation of ART-naive immunosuppressed HIV pts. About 50% of newly diagnosed HIV pts have oral candidiasis.<br>Esophageal candidiasis among advanced HIV pts, not limited to those ART-naive. |
| Fabian 2009(40) | To determine the prevalence of various oral and peri-oral manifestations in people living with HIV/AIDS in Tanzania | Cross sectional study | 187 HIV infected | 45% of participants had at least one oral condition. 32.5% had 2-4 conditions. The conditions in order of frequency: Candidiasis 28.9% (lips/mucosa 22.5%, tongue 18.7%, palate 11.8%, gingiva 10.2%), non- tender lymphadenopathy 11.8%, oral ulcers 12%, angular cheilitis 5.3%, dry mouth 3.7%, oral hairy leukoplakia 3.7%, herpes zoster 3.2%, necrotizing ulcerative gingivitis/ periodontitis 3.2%, Kaposi's sarcoma 0.5%. Malnutrition and low BMI was linked to candidiasis. |
| Boniphace 2011(116) | To share lessons learnt from upscaling ART services | Cross sectional study | 611 HIV care and 284 ART patients | Parotid enlargement not associated with increasing CD4 counts. Esophageal candidiasis was present in 13.1% of subjects. It was more prevalent in low CD4 counts: 61(16.9%) for those with CD4 <100, 16(6.9%) for those with CD4 100-200, and 16 (5.1%) for CD4 >200. |
| Bles 2015(117) | To determine antibiotic susceptibility of colonizing pneumococcal serotypes in HIV-exposed infants before the introduction of the 13-valent pneumococcal conjugate vaccine (PCV13), because HIV-exposed infants are at increased risk of invasive pneumococcal infections | Cross sectional study | 104 pneumococcal isolates | There were no differences in antibiotic susceptibility between vaccine and non-vaccine serotypes. Reduced susceptibility of colonizing pneumococcal isolates for commonly used antibiotics is common in HIV-exposed Tanzanian infants. |
| Anthony 2012(71) | To assess the prevalence of, and risk factors for, pneumococcal carriage in HIV-positive children aged <15 years. | Cross sectional study | 142 HIV-positive children aged <15 years | Eighty one percent of the HIV children had pneumonia in nasopharyngeal swabs. |
| Shija 2020(41) | To identify the magnitude of ENT manifestations among HIV-infected patients attended HIV clinics at KCMC based on age, sex, and CD4 count. | Cross sectional study | HIV-infected patients who attended HIV clinics | Sixty-eight (34%) of 200 HIV-infected patients had ENT manifestations. The most affected age group was 0-9 years. Those with CD4 count less than 200cells/μL also it was a high prevalence (56.3%).<br>13/68 had rhinosinusitis, 11/68 had tonsillitis, 6/68 had esophageal candidiasis, 6/68 had cervical lymphadenopathy, 3/68 had oral candidiasis.<br>We therefore recommend that all clients seeking care at HIV-clinics, particularly those with CD4 counts below 500 cells/μL be screened for ENT conditions. These conditions in this study were not their presenting complaints and were uncovered after screening. |
| Schiødt 1990(42) | To report a detailed study on oral lesions and their association with AIDS criteria and serologic signs of HIV infection in Tanzania | Cross sectional study | 39 hospitalized suspected AIDS patients, 44 I.M. unsuspected patients, 53 dental outpatients, and 50 patients with sexually transmitted diseases. | Oral lesions were found in 54% of those with AIDS criteria and 52% of HIV-infected patients, as compared to 3% and 6% of the patients without AIDS criteria and HIV infection, respectively.<br>Among patients with AIDS atrophic candidiasis occurred in 21% pseudomembranous candidiasis in 23%. hairy leukoplakia in 36%, herpetic stomatitis in 2%, Kaposi's<br>sarcoma in 4%, and nonspecific ulcer in 4%. The presence of oral lesions had a high predictive value for presence of AIDS criteria as well as for presence of HIV infection in this hospital setting. All patients should have a thorough oral examination, and the presence of the oral lesions should lead to testing for HIV infection. |
| Scheutz 1997(118) | To determine whether there is an association between carriage of oral yeasts, malnutrition and HIV-1 infection among Tanzanian children. | Case control study | 882 children between 1.5-5yrs | The presence of HIV predisposes the carriage of oral yeasts in the tongue and buccal regions. |
| Plantinga 2010(119) | To investigate whether genetic variations in the innate immune genes DECTIN-1, TLR2, TLR4, TIRAP, and CASPASE-12 are associated with the presence of OPC in HIV-infected subjects from East Africa. | Cross sectional study | 225 HIV patients | No statistical differences in polymorphism frequencies in the genes. |
| Panya 2009(52) | To determine the prevalence and pattern of mucocutaneous disorders among HIV infected children attending public pediatric 'Care and | Cross sectional study | To determine the prevalence and pattern of mucocutaneous disorders among HIV infected children attending public pediatric 'Care and | 4% oral candidiasis in HIV children. |

|  | Treatment Centres' in Dar es Salaam. |  | Treatment Centres' in Dar es Salaam. |  |
| --- | --- | --- | --- | --- |
| Kisangau 2007(72) | To present the first detailed account of the status and use of traditional medicines in the management of HIV/AIDS opportunistic infections in Tanzania. | Cross sectional study | 30 herbal practitioners | 25% of plant species are used to treat candidiasis. |
| Kibonde 2018(81) | To identify medicinal plant species used to manage HIV/AIDS opportunistic infections by the communities in Rungwe District, Tanzania | Cross sectional study | 193 PLHIV | Oral candidiasis utilized 35% of the species (known medicinal plants) in the treatment |
| Hamza 2008(85) | To compare the clinical and mycological responses, relapse rates, and safety of a single 750-mg dose and a 14-day course of treatment with fluconazole | Randomized controlled trial | 220 HIV-infected patients with clinical and mycological evidence of oropharyngeal candidiasis. | Single-dose and 14-day course fluconazole were both effective in achieving clinical and mycological cure of oropharyngeal candidiasis among HIV patients. |
| Marealle 2021(82) | To document medicinal plants used in the management of HIV and AIDS-related conditions in Mbulu District. | Qualitative research | 6 traditional health professionals and 37 species | 66.7% reported encountering oral thrush.<br>24% of the herbal medicines reported to treat oral candidiasis. |
| Tuominen 1992(92) | To evaluate possible changes in the knowledge of HIV infection and AIDS in | Cross sectional study |  | Dental officers and other members of the operating team were not well aware of the oral manifestations of HIV. |
|  | Tanzanian dental staff after 1 year |  | 89 dental officers, assistant dental officers, and dental assistants | Some could not mention a single oral symptom. |
| Simon 2001(90) | To investigate the types and magnitude of post-extraction complication | Cross sectional study | Dental patients who had their teeth extracted at the Muhimbili Medical Centre dental outpatient clinic | Post-extraction complications are few, mostly minor, self-limiting, and easily treatable. The study does not support routine antibiotic prophylaxis or special pre-extraction procedures, even in this patient population with poor oral hygiene and high HIV seroprevalence. |
| Scheutz 1997(61) | To study the natural association between periodontal condition and HIV infection and the stage of infection | Randomized controlled trial | 119 HIV-infected adults, 73 individuals with AIDS and 156 control HIV negative | No significance in bleeding on probing, pocket formation or attachment loss among the three groups. No significance of the groups to CD4 counts. |
| Matee 1999(91) | To determine the frequency of HIV infection among dental patients attending the three dental facilities at Muhimbili Medical Centre (MMC) in Dar-es-Salaam, Tanzania, and to compare the dental treatment demands and needs of the patients found to be HIV-infected with those of their HIV-seronegative counterpart | Cross sectional study | 460 dental patients | Dental demands/conditions not different between HIV and non-HIV. The high frequency of HIV infection calls for the institution of infection control measures in the dental clinics. |
| Mwakigonja 2007(76) | To elucidate viral and cell biological aspects of oral K.S. to allow formulation of preventive and therapeutic strategies | Cohort study | 74 of 78 subjects were HIV infected | 78 (11%) oral cases out of 700 KS registered, where 69.7% were HIV positive and 112 were unknown. Oral Kaposi Sarcoma is highly associated with HIV and advanced/nodular histological stage.<br>HHV-8 lesional content (LANA immunoreactivity) is higher in nodular oral AIDS-related KS (AKS) lesions of males than females and of children than adults.<br>Males appeared to have more tumor burden (multicentricity and systemization) although OKS frequency among females is seemingly higher.<br>31/78 showed very low CD4 counts. Cell counts of available for (31) OAKS patient blood showed that 25 (80.7%) had CD4 values of less than 50cells/. Concordantly, most patients (24/31) (77.4%) had a CD4/CD8 ratio <0.1 implying severe immune deficiency. |
| Gilyoma 2015(78) | To describe the clinicopathological profile of HNC in our local setting and highlights the challenges in the management of this disease. | Cohort study | 346 confirmed cases of histologically confirmed H.N. cancers | No statistically significance between HAART and HNC. |
| Campbell 2016(74) | To gain further understanding of Tanzania's changing cancer burden | Cross sectional study | 1127 patients diagnosed/treated in 2010-2014 plus all ORCI patients diagnosed/treated for lung, liver, and HNC 2002-2014 =2067 | 7.5% of all head and neck cancer cases were HIV-positive. 81.5% of HIV patients with NADC had head and neck cancer. Of these 81.5, 44.4% of HIV-positive patients were diagnosed with cancer of the oral cavity/oropharynx, 27.2% had cancer of the nasal cavity/paranasal sinuses, and 9.9% had cancer of the hypopharynx/ larynx/ trachea.<br>Lastly, over half the patients (HIV plus HNC) diagnosed with these NADCs during 2010-2014 presented to ORCI at advanced stages of malignancy. |
| Koski 2015(45) | To examine the trend and possible changes in annual proportions of K.S. in relation to all cancers over a 10-year period (2002-2011) that correlated with increased access to ARTs and examine the sociodemographic, clinical, and treatment characteristics of patients with K.S. during the study period | Cross sectional study | 1504 KS from ORCI, of which 72.5% were HIV positive | ART duration showed a protective effect with oral lesions (OR=0.55, CI: 0.33, 0.91). |

## Appendix: Search Strategies

### MEDLINE (via PubMed)

**Search date: 2/9/2024**

| Concept | Strategy | Results |
| --- | --- | --- |
| #1 Oral Health / Cancer | "Oral health"[Mesh] OR "Mouth Diseases"[Mesh] OR "Oropharynx"[Mesh] OR "Parotid Diseases"[Mesh] OR "Salivary Glands"[Mesh] OR "Pharynx"[Mesh] OR "Larynx"[Mesh] OR "Tongue"[Mesh] OR "Lip"[Mesh] OR "Nasopharyngeal Neoplasms"[Mesh] OR "Nasopharyngeal Carcinoma"[Mesh] OR "Oropharyngeal Neoplasms"[Mesh] OR "Hypopharyngeal Neoplasms"[Mesh] OR oral[tiab] OR mouth[tiab] OR mouths[tiab] OR "oropharynx"[tiab] OR "parotid diseases"[tiab] OR "parotid disease"[tiab] OR dentition[tiab] OR mouth[tiab] OR mouths[tiab] OR "salivary glands"[tiab] OR "salivary gland"[tiab] OR "salivary duct"[tiab] OR "salivary ducts"[tiab] OR tongue[tiab] OR tongues[tiab] OR "Pharynx"[tiab] OR "Pharynx"[tiab] OR "throat"[tiab] OR "throats"[tiab] OR "Nasopharyngeal Neoplasms"[Tiab] OR "Nasopharyngeal Carcinoma"[Tiab] OR "Oropharyngeal Neoplasms"[Tiab] OR "Hypopharyngeal Neoplasms"[Tiab] OR larynx[tiab] OR sinonasal[tiab] OR sino-nasal[tiab] | 1,286,368 |
| #2 HPV | "Human Papillomavirus Viruses"[Mesh] OR "Human papillomavirus 11"[Mesh] OR "Human papillomavirus 16"[Mesh] OR "Human papillomavirus 18"[Mesh] OR "Human papillomavirus 31"[Mesh] OR "Human papillomavirus 6"[Mesh] OR "Human Papillomavirus Virus"[tiab] OR "Human Papillomavirus Viruses" OR HPV[tiab] OR "Human Papillomavirus Recombinant Vaccine Quadrivalent, Types 6, 11, 16, 18"[Mesh] OR "Papillomavirus Vaccines"[Mesh] OR Gardasil[tiab] OR "HPV vaccine"[tiab] OR "HPV vaccines"[tiab] OR "human papilloma virus vaccine"[tiab] OR "human papilloma virus vaccines"[tiab] OR "papillomavirus vaccine"[tiab] OR "papillomavirus vaccines"[tiab] OR "HPV immunization"[tiab] OR "papilloma virus immunization"[tiab] OR "papillomavirus immunization"[tiab] OR "HPV immunizations"[tiab] OR "papilloma virus immunizations"[tiab] OR "papillomavirus immunizations"[tiab] OR "Cervarix"[tiab] OR GAVI[tiab] OR "Global Alliance for Vaccines and Immunization"[tiab] | 57,265 |
| #3 HIV | "HIV infections"[Mesh] OR "HIV"[Mesh] OR HIV[tiab] OR "Acquired Immunodeficiency Syndrome"[Mesh] OR "human immunodeficiency virus"[tiab] OR AIDS[tiab] OR "acquired immune deficiency syndrome"[tiab] OR "Acquired Immuno-Deficiency Syndrome"[tiab] OR "Acquired Immuno Deficiency Syndrome"[tiab] OR "Acquired Immunologic Deficiency | 512,814 |
|  | Syndrome"[tiab] OR "Human T Cell Lymphotropic Virus"[tiab] OR "Human T Cell Leukemia Virus"[tiab] OR "LAV-HTLV-III"[tiab] OR "Lymphadenopathy-Associated Virus"[tiab] OR "Lymphadenopathy Associated Virus"[tiab] OR "Lymphadenopathy-Associated Viruses"[tiab] |  |
| #4 Tanzania | "Tanzania"[Mesh] OR "Tanzania"[all fields] OR "United Republic of Tanzania"[all fields] OR "Zanzibar"[all fields] OR "Tanganyika"[all fields] OR "Tanzanian"[all fields] OR "Dodoma"[all fields] OR "Dar es Salaam"[all fields] | 25,769 |
| #5 Combined HPV OR HIV | #2 OR #3 | 566,063 |
| #6 Combine all | #1 AND #4 AND #5 | 194 |
| # Validation String | **10858748 OR 10702788 OR 25875349 OR 16916469 OR 18694525 OR 18840077 OR 19774801** | 7/7 |

### Embase (via Elsevier)

**Search date: 2/9/2024**

Embase (via Elsevier) Search date: 2/9/2024
| Concept | Strategy | Results |
| --- | --- | --- |
| #1 Oral Health / Cancer | 'mouth disease'/exp OR 'oropharynx'/exp OR 'parotid gland disease'/exp OR 'salivary gland'/exp OR 'pharynx'/exp OR 'pharynx disease'/exp OR 'throat disease'/exp OR 'larynx'/exp OR 'larynx disorder'/exp OR 'tongue'/exp OR 'tongue disease'/exp OR 'nasopharynx tumor'/exp OR 'oropharynx tumor'/exp OR 'hypopharynx tumor'/exp OR (oral OR mouth OR mouths OR 'oropharynx' OR 'parotid diseases' OR 'parotid disease' OR dentition OR mouth OR mouths OR 'salivary glands' OR 'salivary gland' OR 'salivary duct' OR 'salivary ducts' OR tongue OR tongues OR 'Pharynx' OR 'Pharynxes' OR 'throat' OR 'throats' OR 'Nasopharyngeal Neoplasms' OR 'Nasopharyngeal Carcinoma' OR 'Oropharyngeal Neoplasms' OR 'Hypopharyngeal Neoplasms' OR larynx OR sinonasal OR sino-nasal):ti,ab | 2,052,699 |
| #2 HPV | 'Wart virus'/exp OR 'Human papillomavirus type 11'/exp OR 'Human papillomavirus type 16'/exp OR 'Human papillomavirus type 18'/exp OR 'Human papillomavirus type 31'/exp OR 'Human papillomavirus type 6'/exp OR ('Human Papillomavirus Virus' OR 'Human Papillomavirus Viruses' OR HPV OR Gardasil OR 'HPV vaccine' OR 'HPV vaccines' OR 'human papilloma virus vaccine' OR 'human papilloma virus vaccines' OR 'papillomavirus vaccine' OR 'papillomavirus vaccines' OR 'HPV immunization' OR 'papilloma virus immunization' OR 'papillomavirus immunization' OR 'HPV immunizations' OR 'papilloma virus immunizations' OR 'papillomavirus immunizations' OR 'Cervarix' | 90,322 |
|  | OR GAVI OR 'Global Alliance for Vaccines and Immunization'):ti,ab |  |
| #3 HIV | 'Human immunodeficiency virus infection'/exp OR 'Human immunodeficiency virus'/exp OR 'acquired immune deficiency syndrome'/exp OR ('human immunodeficiency virus' OR AIDS OR 'acquired immune deficiency syndrome' OR 'Acquired Immuno-Deficiency Syndrome' OR 'Acquired Immuno Deficiency Syndrome' OR 'Acquired Immunologic Deficiency Syndrome' OR 'Human T Cell Lymphotropic Virus' OR 'Human T Cell Leukemia Virus' OR 'LAV-HTLV-III' OR 'Lymphadenopathy-Associated Virus' OR 'Lymphadenopathy Associated Virus' OR 'Lymphadenopathy-Associated Viruses'):ti,ab | 1,057,097 |
| #4 Tanzania | 'Tanzania'/exp OR ('Tanzania' OR 'United Republic of Tanzania' OR 'Zanzibar' OR 'Tanganyika' OR 'Tanzanian' OR 'Dodoma' OR 'Dar es Salaam'):ti,ab,ff,ad,ca | 32,017 |
| #5<br>Combined<br>HPV OR<br>HIV | #2 OR #3 | 1,139,690 |
| #6 Combine<br>all | #1 AND #4 AND #5 | 327 |

### Web of Science (via Clarivate)

**Search date: 2/9/2024**

Web of Science (via Clarivate) Search date: 2/9/2024
| Concept | Strategy | Results |
| --- | --- | --- |
| #1 Oral<br>Health /<br>Cancer | TS=(oral OR mouth OR mouths OR "oropharynx" OR "parotid diseases" OR "parotid disease" OR dentition OR mouth OR mouths OR "salivary glands" OR "salivary gland" OR "salivary duct" OR "salivary ducts" OR tongue OR tongues OR "Pharynx" OR "Pharynx" OR "throat" OR "throats" OR "Nasopharyngeal Neoplasms" OR "Nasopharyngeal Carcinoma" OR "Oropharyngeal Neoplasms" OR "Hypopharyngeal Neoplasms" OR larynx OR sinonasal OR sino-nasal) | 1,077,356 |
| #2 HPV | TS=("Human Papillomavirus Virus" OR "Human Papillomavirus Viruses" OR HPV OR Gardasil OR "HPV vaccine" OR "HPV vaccines" OR "human papilloma virus vaccine" OR "human papilloma virus vaccines" OR "papillomavirus vaccine" OR "papillomavirus vaccines" OR "HPV immunization" OR "papilloma virus immunization" OR "papillomavirus immunization" OR "HPV immunizations" OR "papilloma virus immunizations" OR "papillomavirus immunizations" OR "Cervarix" OR GAVI OR "Global Alliance for Vaccines and Immunization") | 60,618 |
| #3 HIV | TS=("human immunodeficiency virus" OR AIDS OR "acquired immune deficiency syndrome" OR "Acquired Immuno- | 861,352 |
|  | Deficiency Syndrome" OR "Acquired Immuno Deficiency Syndrome" OR "Acquired Immunologic Deficiency Syndrome" OR "Human T Cell Lymphotropic Virus" OR "Human T Cell Leukemia Virus" OR "LAV-HTLV-III" OR "Lymphadenopathy-Associated Virus" OR "Lymphadenopathy Associated Virus" OR "Lymphadenopathy-Associated Viruses") |  |
| #4 Tanzania | TS=("Tanzania" OR "United Republic of Tanzania" OR "Zanzibar" OR "Tanganyika" OR "Tanzanian" OR "Dodoma" OR "Dar es Salaam") OR OG=("Tanzania" OR "United Republic of Tanzania" OR "Zanzibar" OR "Tanganyika" OR "Tanzanian" OR "Dodoma" OR "Dar es Salaam") OR AD=("Tanzania" OR "United Republic of Tanzania" OR "Zanzibar" OR "Tanganyika" OR "Tanzanian" OR "Dodoma" OR "Dar es Salaam") | 55,276 |
| #5 Combined HPV OR HIV | #2 OR #3 | 919,259 |
| #6 Combine all | #1 AND #4 AND #5 | 95 |

### Global Index Medicus (via World Health Organization)

**Search date: 2/9/2024**

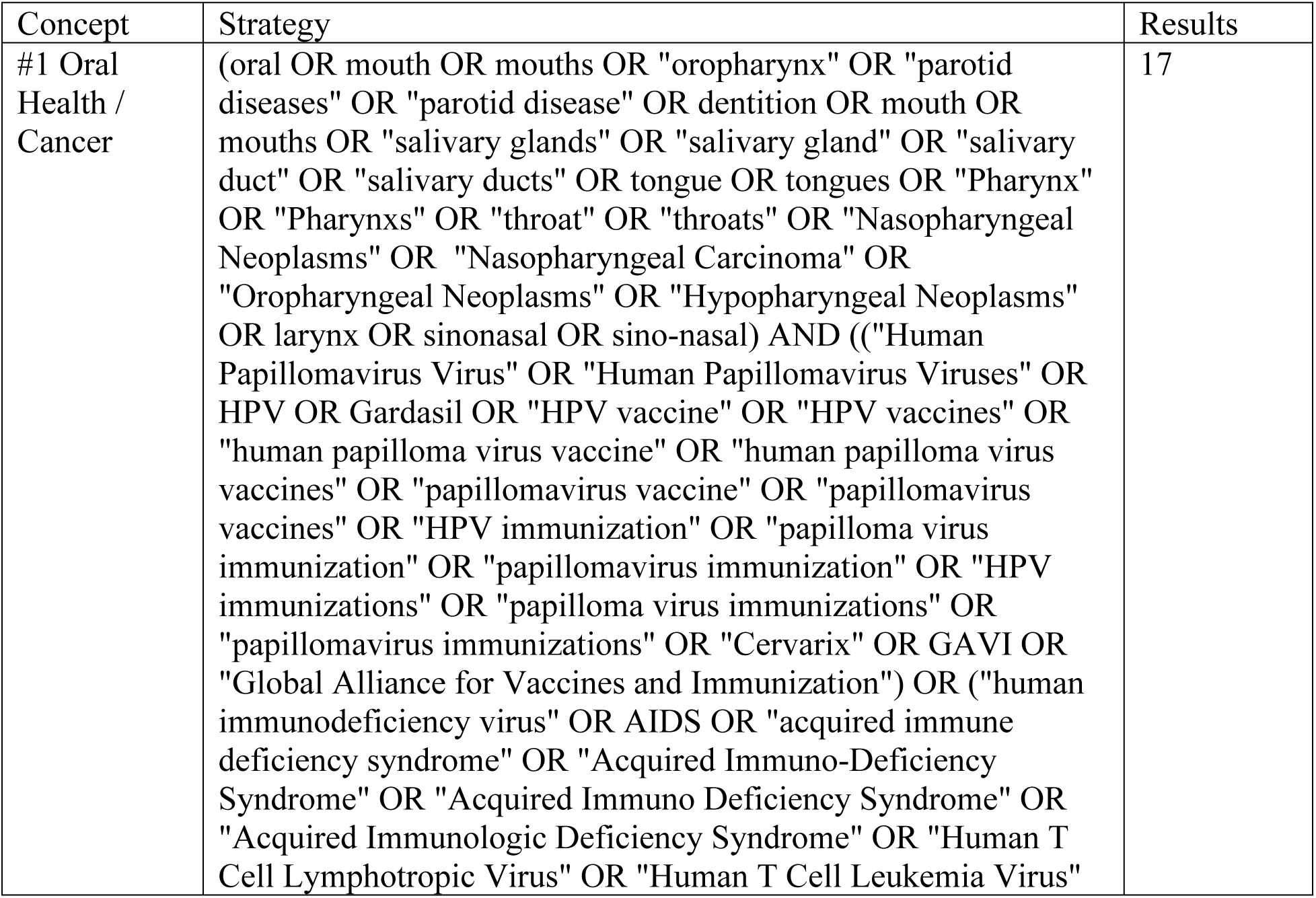
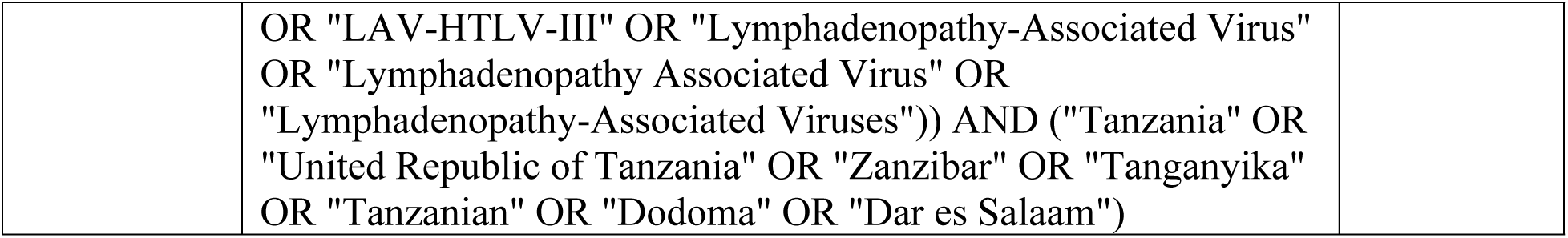

### CAB Global Health (via EBSCOhost)

**Search date: 2/9/2024**

CAB Global Health (via EBSCOhost) Search date: 2/9/2024
| Concept | Strategy | Results |
| --- | --- | --- |
| S1 Oral Health / Cancer | DE "mouth diseases" OR DE "noma" OR DE "oral cancer" OR DE "oral hairy leukoplakia" OR DE "periodontal diseases" OR DE "salivary gland diseases" OR DE "stomatitis" OR DE "tongue diseases" OR DE "tooth diseases" OR DE "salivary glands" OR DE "mandibular glands" OR DE "parotid gland" OR DE "pharynx" OR DE "nasopharynx" OR DE "larynx" OR DE "tongue" OR DE "tongue cancer" OR DE "tongue diseases" OR DE "glossitis" OR DE "nasopharynx" OR TI oral OR TI mouth OR TI mouths OR TI "oropharynx" OR TI "parotid diseases" OR TI "parotid disease" OR TI dentition OR TI mouth OR TI mouths OR TI "salivary glands" OR TI "salivary gland" OR TI "salivary duct" OR TI "salivary ducts" OR TI tongue OR TI tongues OR TI "Pharynx" OR TI "Pharynxes" OR TI "throat" OR TI "throats" OR TI "Nasopharyngeal Neoplasms" OR TI "Nasopharyngeal Carcinoma" OR TI "Oropharyngeal Neoplasms" OR TI "Hypopharyngeal Neoplasms" OR TI larynx OR TI sinonasal OR TI sino-nasal OR AB oral OR AB mouth OR AB mouths OR AB "oropharynx" OR AB "parotid diseases" OR AB "parotid disease" OR AB dentition OR AB mouth OR AB mouths OR AB "salivary glands" OR AB "salivary gland" OR AB "salivary duct" OR AB "salivary ducts" OR AB tongue OR AB tongues OR AB "Pharynx" OR AB "Pharynxes" OR AB "throat" OR AB "throats" OR AB "Nasopharyngeal Neoplasms" OR AB "Nasopharyngeal Carcinoma" OR AB "Oropharyngeal Neoplasms" OR AB "Hypopharyngeal Neoplasms" OR AB larynx OR AB sinonasal OR AB sino-nasal | 250,514 |
| S2 HPV | DE "Human papillomavirus 18" OR DE "Human papillomavirus 6" OR TI "Human Papillomavirus Virus" OR TI "Human Papillomavirus Viruses" OR TI HPV OR TI Gardasil OR TI "HPV vaccine" OR TI "HPV vaccines" OR TI "human papilloma virus vaccine" OR TI "human papilloma virus vaccines" OR TI "papillomavirus vaccine" OR TI "papillomavirus vaccines" OR TI "HPV immunization" OR TI "papilloma virus immunization" OR TI "papillomavirus immunization" OR TI "HPV immunizations" OR TI "papilloma virus immunizations" OR TI "papillomavirus immunizations" OR TI "Cervarix" OR TI GAVI OR TI "Global Alliance for Vaccines and Immunization" OR AB | 22,276 |
|  | "Human Papillomavirus Virus" OR AB "Human Papillomavirus Viruses" OR AB HPV OR AB Gardasil OR AB "HPV vaccine" OR AB "HPV vaccines" OR AB "human papilloma virus vaccine" OR AB "human papilloma virus vaccines" OR AB "papillomavirus vaccine" OR AB "papillomavirus vaccines" OR AB "HPV immunization" OR AB "papilloma virus immunization" OR AB "papillomavirus immunization" OR AB "HPV immunizations" OR AB "papilloma virus immunizations" OR AB "papillomavirus immunizations" OR AB "Cervarix" OR AB GAVI OR AB "Global Alliance for Vaccines and Immunization" |  |
| S3 HIV | DE "HIV infections" OR DE "HIV-1 infections" OR DE "HIV-2 infections" OR DE "HIV-1 infections" OR DE "HIV-2 infections" OR DE "human immunodeficiency viruses" OR DE "Human immunodeficiency virus 1" OR DE "Human immunodeficiency virus 2" OR DE "acquired immune deficiency syndrome" OR TI "human immunodeficiency virus" OR TI AIDS OR TI "acquired immune deficiency syndrome" OR TI "Acquired Immuno-Deficiency Syndrome" OR TI "Acquired Immuno Deficiency Syndrome" OR TI "Acquired Immunologic Deficiency Syndrome" OR TI "Human T Cell Lymphotropic Virus" OR TI "Human T Cell Leukemia Virus" OR TI "LAV-HTLV-III" OR TI "Lymphadenopathy-Associated Virus" OR TI "Lymphadenopathy Associated Virus" OR TI "Lymphadenopathy-Associated Viruses" OR AB "human immunodeficiency virus" OR AB AIDS OR AB "acquired immune deficiency syndrome" OR AB "Acquired Immuno-Deficiency Syndrome" OR AB "Acquired Immuno Deficiency Syndrome" OR AB "Acquired Immunologic Deficiency Syndrome" OR AB "Human T Cell Lymphotropic Virus" OR AB "Human T Cell Leukemia Virus" OR AB "LAV-HTLV-III" OR AB "Lymphadenopathy-Associated Virus" OR AB "Lymphadenopathy Associated Virus" OR AB "Lymphadenopathy-Associated Viruses" | 220,476 |
| S4 Tanzania | DE "Tanzania" OR DE "Zanzibar" OR TI "Tanzania" OR TI "United Republic of Tanzania" OR TI "Zanzibar" OR TI "Tanganyika" OR TI "Tanzanian" OR TI "Dodoma" OR TI "Dar es Salaam" OR AB "Tanzania" OR AB "United Republic of Tanzania" OR AB "Zanzibar" OR AB "Tanganyika" OR AB "Tanzanian" OR AB "Dodoma" OR AB "Dar es Salaam" | 17,565 |
| S5 Combined HPV OR HIV | S2 OR S3 | 240,728 |
| S6 Combine all | S1 AND S4 AND S5 | 12312 |

((“Oral health” OR “Mouth Diseases” OR “Oropharynx” OR “Parotid Diseases” OR “Salivary Glands” OR “Pharynx” OR “Larynx” OR “Tongue” OR “Lip” OR “Nasopharyngeal Neoplasms” OR “Nasopharyngeal Carcinoma” OR “Oropharyngeal Neoplasms” OR “Hypopharyngeal Neoplasms” OR oral OR mouth OR mouths OR “oropharynx” OR “parotid diseases” OR “parotid disease” OR dentition OR mouth OR mouths OR “salivary glands” OR “salivary gland” OR “salivary duct” OR “salivary ducts” OR tongue OR tongues OR “Pharynx” OR “Pharynxs” OR “throat” OR “throats” OR “Nasopharyngeal Neoplasms” OR “Nasopharyngeal Carcinoma” OR “Oropharyngeal Neoplasms” OR “Hypopharyngeal Neoplasms” OR larynx OR sinonasal OR sino-nasal) AND (“Human Papillomavirus Viruses” OR “Human papillomavirus 11” OR “Human papillomavirus 16” OR “Human papillomavirus 18” OR “Human papillomavirus 31” OR “Human papillomavirus 6” OR “Human Papillomavirus Virus” OR “Human Papillomavirus Viruses” OR HPV OR “Human Papillomavirus Recombinant Vaccine Quadrivalent, Types 6, 11, 16, 18” OR “Papillomavirus Vaccines” OR Gardasil OR “HPV vaccine” OR “HPV vaccines” OR “human papilloma virus vaccine” OR “human papilloma virus vaccines” OR “papillomavirus vaccine” OR “papillomavirus vaccines” OR “HPV immunization” OR “papilloma virus immunization” OR “papillomavirus immunization” OR “HPV immunizations” OR “papilloma virus immunizations” OR “papillomavirus immunizations” OR “Cervarix” OR GAVI OR “Global Alliance for Vaccines and Immunization”)) AND (“HIV infections” OR “HIV” OR HIV OR “Acquired Immunodeficiency Syndrome” OR “human immunodeficiency virus” OR AIDS OR “acquired immune deficiency syndrome” OR “Acquired Immuno-Deficiency Syndrome” OR “Acquired Immuno Deficiency Syndrome” OR “Acquired Immunologic Deficiency Syndrome” OR “Human T Cell Lymphotropic Virus” OR “Human T Cell Leukemia Virus” OR “LAV-HTLV-III” OR “Lymphadenopathy-Associated Virus” OR “Lymphadenopathy Associated Virus” OR “Lymphadenopathy-Associated Viruses”)

